# Clinical Characteristics and Outcomes of COVID-19 patients with diabetes mellitus in Kuwait

**DOI:** 10.1101/2020.08.20.20178525

**Authors:** Abdullah Alshukry, Mohammad Bu Abbas, Yaseen Ali, Barrak Alahmad, Abdullah A. Al-Shammari, Ghadeer Alhamar, Mohammad Abu-Farha, Jehad AbuBaker, Sriraman Devarajan, Ali A. Dashti, Fahd Al-Mulla, Hamad Ali

## Abstract

**Background:** COVID-19 has a highly variable clinical presentation, ranging from asymptomatic to severe respiratory symptoms and death. Diabetes seems to be one of the main comorbidities contributing to a worse COVID-19 outcome.

**Objective:** In here we analyze the clinical characteristics and outcomes of diabetic COVID-19 patients Kuwait.

**Methods:** In this single-center, retrospective study of 417 consecutive COVID-19 patients, we analyze and compare disease severity, outcome, associated complications, and clinical laboratory findings between diabetic and non-diabetic COVID-19 patients.

**Results:** COVID-19 patients with diabetes had more ICU admission than non-diabetic COVID-19 patients (20.1% vs. 16.8%, p<0.001). Diabetic COVID-19 patients also recorded higher mortality in comparison to non-diabetic COVID-19 patients (16.7% vs. 12.1%, p<0.001). Diabetic COVID-19 patients had significantly higher prevalence of comorbidities, such as hypertension. Laboratory investigations also highlighted notably higher levels of C-reactive protein in diabetic COVID019 patients and lower estimated glomerular filtration rate. They also showed a higher incidence of complications. logistic regression analysis showed that every 1 mmol/L increase in fasting blood glucose in COVID-19 patients is associated with 1.52 (95% CI: 1.34 – 1.72, p<0.001) times the odds of dying from COVID-19.

**Conclusion:** Diabetes is a major contributor to worsening outcomes in COVID-19 patients. Understanding the pathophysiology underlining these findings could provide insight into better management and improved outcome of such cases.

**Highlights of the Study:**

- A significantly higher proportion of COVID-19 patients with diabetes mellitus required admission to the ICU.
- Higher fasting blood glucose was associated with higher risk of COVID-19 associated mortality.
- COVID-19 patients with diabetes mellitus had significantly higher incidence of complications including sepsis, ARDS, cardiac failure and renal failure.

## 1. Introduction

In December 2019, a novel coronavirus, now known as Severe Acute Respiratory Syndrome Coronavirus 2 (SARS-CoV-2), emerged in Wuhan, China^1-3^. Since then, it has spread rapidly all over the world and was declared a pandemic by the World Health Organization on March 11, 2020. With the number of cases currently exeeding 100 million and more than two million deaths, the virus poses a major threat to global health^4,5^.

Coronavirus Disease 2019 (COVID-19), the disease caused by the virus, has shown a highly variable clinical presentation, ranging from asymptomatic to severe illness leading to death. The symptoms of COVID-19 include fever, cough, dyspnea, myalgia, fatigue, headache, and loss of taste or smell. Most patients experience mild symptoms, although some may develop serious complications, including acute respiratory distress syndrome (ARDS), multiorgan failure, septic shock, and hypercoagulation, which can eventually lead to death^6-10^. The exact reasons for the observed variability in disease manifestations and outcomes are not fully understood. Whereas pediatric cases show a milder clinical course^11^, a worse prognosis has been associated with older age and being male. Emerging evidence also indicates that preexisting medical conditions, including hypertension, cardiovascular disease, chronic kidney disease, chronic obstructive pulmonary disease, solid organ transplantation, and diabetes, can increase the risk of poor COVID-19 prognosis^2,12-17^.

Diabetes mellitus is a chronic metabolic disease characterized by the occurrence of hyperglycemia for a prolonged period. It is associated with serious long-term complications, including cardiovascular disease and chronic kidney disease ^18^. Diabetes, which affects 463 million people, has a major impact on global health. It is among the top 10 causes of adult deaths worldwide^19^. In the past two decades, two coronaviruses have emerged that have caused widespread respiratory illness and deaths. In 2002, SARS coronavirus (SARS-CoV) emerged in China, causing severe acute respiratory syndrome coronavirus. In 2012, another coronavirus, MERS-coronavirus (MERS-CoV), emerged in Saudi Arabia, causing Middle East respiratory syndrome. Experience with both syndromes revealed that diabetes was a risk factor for poor prognosis and mortality^20,21^. Similarly, initial reports of the clinical characteristics of COVID-19 showed a similar trend^1,22^. Given the global burden of diabetes and the pandemic course of SARS-CoV-2, understanding how diabetes contributes to a worse COVID-19 prognosis is important.

Several studies have shown that type 2 diabetes, which is the most prevalent type of the disease, is associated with low-grade chronic inflammation that affects the homeostatic glucose regulation and insulin sensitivity^23,24^. Studies have indicated that hyperglycemia may be a major pushing factor for a poorer prognosis of COVID-19, and has been a suggested risk factor for predicting ICU admission and negative outcomes in patients ^17^. This finding has been seen in earlier SARs-CoV infections, where hyperglycemia and presence of diabetes were independent predictors of mortality^21^. Furthermore, hyperglycemia has been indicative of a poorer prognosis of COVID-19, regardless of diabetes status^17^. Hyperglycemia in combination with chronic inflammation could contribute to an abnormal immune response by weakening T-cell function, in addition to an increased risk of hyperinflammation and cytokine storm^25,26^, which in turn can worsen the COVID-19 disease outcome.

Understanding how diabetes worsens COVID-19 outcomes can help provide better disease management and contribute to the improvement in disease outcomes. In this study, we perform a comprehensive clinical analysis of COVID-19 patients with and without type 2 diabetes. We analyze and compare the distribution of disease severity, associated complications, and death outcomes between the two groups.

## 2. Subjects and Methods

### 2.1 Study design

The Standing Committee for Coordination of Health and Medical Research at the Ministry of Health in Kuwait reviewed and approved this retrospective study (Institutional Review Board 2020/1404). The Standing Committee waived the requirement for written informed consent because of the urgency of data collection and the exceptional nature of the disease. All procedures involving human participants were performed following the relevant guidelines and regulations. The medical records of confirmed COVID-19 cases admitted to Jaber Al-Ahmad Hospital in Kuwait between February 24 and May 24, 2020, were accessed, analyzed, and included in this study. The diagnosis of COVID-19 was established based on positive viral real-time reverse transcriptase-polymerase chain reaction (RT-PCR) assay of nasal and/or pharyngeal swabs, following the World Health Organization’s interim guidance. Cases were divided into two main groups: patients with diabetes and patients without diabetes. The diagnosis of diabetes was based on a fasting plasma glucose value of ≥7.0 mmol/L and/or patient’s medical history ^27^. Only type 2 diabetes patients were considered in this study as no type 1 diabetes patients were encountered. Laboratory diagnosis was confirmed with medical history. Each group was further divided into the following subgroups depending on COVID-19 severity and outcome: asymptomatic, symptomatic with mild/moderate symptoms, intensive care unit (ICU) survivors, and ICU death. End points analyzed for comparison between diabetic and non-diabetic patients are ICU admission and death. Data presented in this study was made available on springer nature data depository^28^ and can be accessed via https://doi.org/10.6084/m9.figshare.12567881.v1.

### 2.2 Data collection

We included 417 confirmed COVID-19 patients in the study. All patients were hospitalized in Jaber Al-Ahmad Hospital, a major governmental hospital which was made exclusive for COVID-19 patients. The patients’ medical records were accessed and analyzed by our team at Dasman Diabetes Institute, Faculty of Allied Health Sciences at Kuwait University, and Jaber Al-Ahmad Hospital. We obtained and analyzed demographic data, medical history, including underlying comorbidities, travel history, contact tracing data, clinical chemistry, hematological laboratory findings, chest radiological images, treatments, complications, ICU admissions and durations, and dynamics of hospital stay and outcomes. The diagnosis of ARDS was determined based on the Berlin definition^29^. Acute kidney injury was evaluated following the Kidney Disease: Improving Global Outcomes definition^30^. The presence of cardiac injury was established based on cardiac blood markers, electrocardiography, and/or echocardiography^9^.

### 2.3 Hospitalization dynamics

During the patient recruitment period, Jaber Al-Ahmad Hospital by the Ministry of Health instituted a 100% hospitalization policy for COVID-19–positive cases. All cases with a positive RT-PCR test, including asymptomatic cases, were admitted, isolated, and put under medical surveillance. Patients in the mild/moderate group, who were hemodynamically stable and had no signs of respiratory distress, were admitted to the ward after RT-PCR confirmation for isolation, medical surveillance, and reevaluation. Patients were transferred to the ICU if they developed signs of respiratory distress and desaturation of oxygen levels (confirmed by pulse oximetry and arterial blood gases) and/or signs of hemodynamic instability that required close monitoring and intensive re-establishment of homeostasis. Patients with severe to critical COVID-19 symptoms were admitted directly to the ICU if they matched any of the following criteria of severity: hypoxemic respiratory failure requiring respiratory support, such as patients who developed ARDS; hemodynamic instability due to cardiogenic or septic shock and clinical, radiological, or laboratory evidence of heart failure; acute cardiac injury; and acute kidney injury secondary to COVID-19 manifestations.

### 2.4 Statistical analysis

The variables analyzed in the study were divided into categorical and continuous variables. The categorical variables were described as frequencies and percentages, whereas continuous variables were presented as medians and interquartile ranges and means and standard deviations. We used a one-way analysis of variance to compare means between groups; we used the Kruskal–Wallis H test to compare the medians of the different group laboratory parameters. Categorical variables were analyzed using the chi-square test, and when the data were limited, Fisher exact test was used. The differences between group means and medians were considered statistically significant when *p* < 0.05. We investigated the relationship between fasting blood glucose as a continuous exposure and ICU admission as an outcome from COVID-19 as a binary response and adjusted to other covariates (age, gender, smoking status, diabetes status, and other comorbidities). We employed a logistic regression model and reported the odds ratios of the outcome of dying from COVID-19 for each 1 mmol/L increase in fasting blood glucose. All statistical analyses were performed using GraphPad Prism software (La Jolla, CA, USA), SPSS (Statistical Package for Social Sciences) for Windows version 25.0 (IBM SPSS Inc., Chicago, IL, USA) and R version 3.4.3 (R Foundation for Statistical Computing, Vienna, Austria).

## 3. Results

### 3.1 Cohort characteristics

The studied cohort consisted of 417 COVID-19 patients who were divided into two groups based on fasting blood glucose levels: 273 (65.5%) non-diabetic patients and 122 (29.3%) diabetic patients (Table 1). The mean age of the non-diabetic group was 39.55 (± 16.59) years, whereas the mean age of the diabetic group was 56.44 (± 11.64) years (*p* < 0.001, Student *t*-test; Table 1 and supplementary Figure 1). The diabetic group had a greater incidence of fever, shortness of breath, and fatigue than the non-diabetic group (*p* < 0.05, < 0.001, and < 0.05, respectively). However, the differences in other symptoms were not significant between the two groups (Table 1).

**Table 1.**
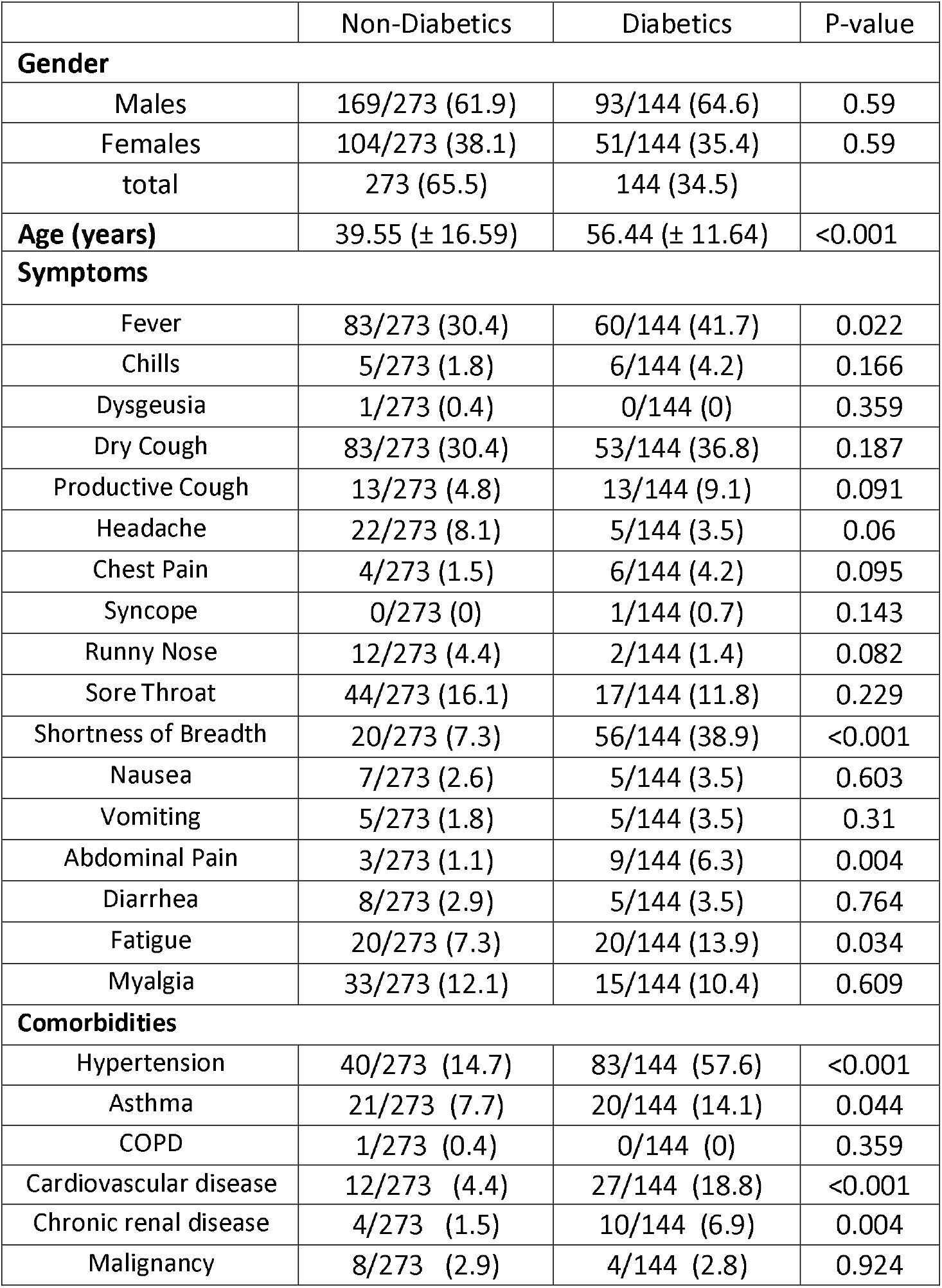
Cohort characteristics stratified by diabetic status

|  | Non-Diabetics | Diabetics | P-value |
| --- | --- | --- | --- |
| **Gender** |  |  |  |
| Males | 169/273 (61.9) | 93/144 (64.6) | 0.59 |
| Females | 104/273 (38.1) | 51/144 (35.4) | 0.59 |
| total | 273 (65.5) | 144 (34.5) |  |
| **Age (years)** | 39.55 (± 16.59) | 56.44 (± 11.64) | <0.001 |
| **Symptoms** |  |  |  |
| Fever | 83/273 (30.4) | 60/144 (41.7) | 0.022 |
| Chills | 5/273 (1.8) | 6/144 (4.2) | 0.166 |
| Dysgeusia | 1/273 (0.4) | 0/144 (0) | 0.359 |
| Dry Cough | 83/273 (30.4) | 53/144 (36.8) | 0.187 |
| Productive Cough | 13/273 (4.8) | 13/144 (9.1) | 0.091 |
| Headache | 22/273 (8.1) | 5/144 (3.5) | 0.06 |
| Chest Pain | 4/273 (1.5) | 6/144 (4.2) | 0.095 |
| Syncope | 0/273 (0) | 1/144 (0.7) | 0.143 |
| Runny Nose | 12/273 (4.4) | 2/144 (1.4) | 0.082 |
| Sore Throat | 44/273 (16.1) | 17/144 (11.8) | 0.229 |
| Shortness of Breadth | 20/273 (7.3) | 56/144 (38.9) | <0.001 |
| Nausea | 7/273 (2.6) | 5/144 (3.5) | 0.603 |
| Vomiting | 5/273 (1.8) | 5/144 (3.5) | 0.31 |
| Abdominal Pain | 3/273 (1.1) | 9/144 (6.3) | 0.004 |
| Diarrhea | 8/273 (2.9) | 5/144 (3.5) | 0.764 |
| Fatigue | 20/273 (7.3) | 20/144 (13.9) | 0.034 |
| Myalgia | 33/273 (12.1) | 15/144 (10.4) | 0.609 |
| **Comorbidities** |  |  |  |
| Hypertension | 40/273 (14.7) | 83/144 (57.6) | <0.001 |
| Asthma | 21/273 (7.7) | 20/144 (14.1) | 0.044 |
| COPD | 1/273 (0.4) | 0/144 (0) | 0.359 |
| Cardiovascular disease | 12/273 (4.4) | 27/144 (18.8) | <0.001 |
| Chronic renal disease | 4/273 (1.5) | 10/144 (6.9) | 0.004 |
| Malignancy | 8/273 (2.9) | 4/144 (2.8) | 0.924 |

**Figure 1.**
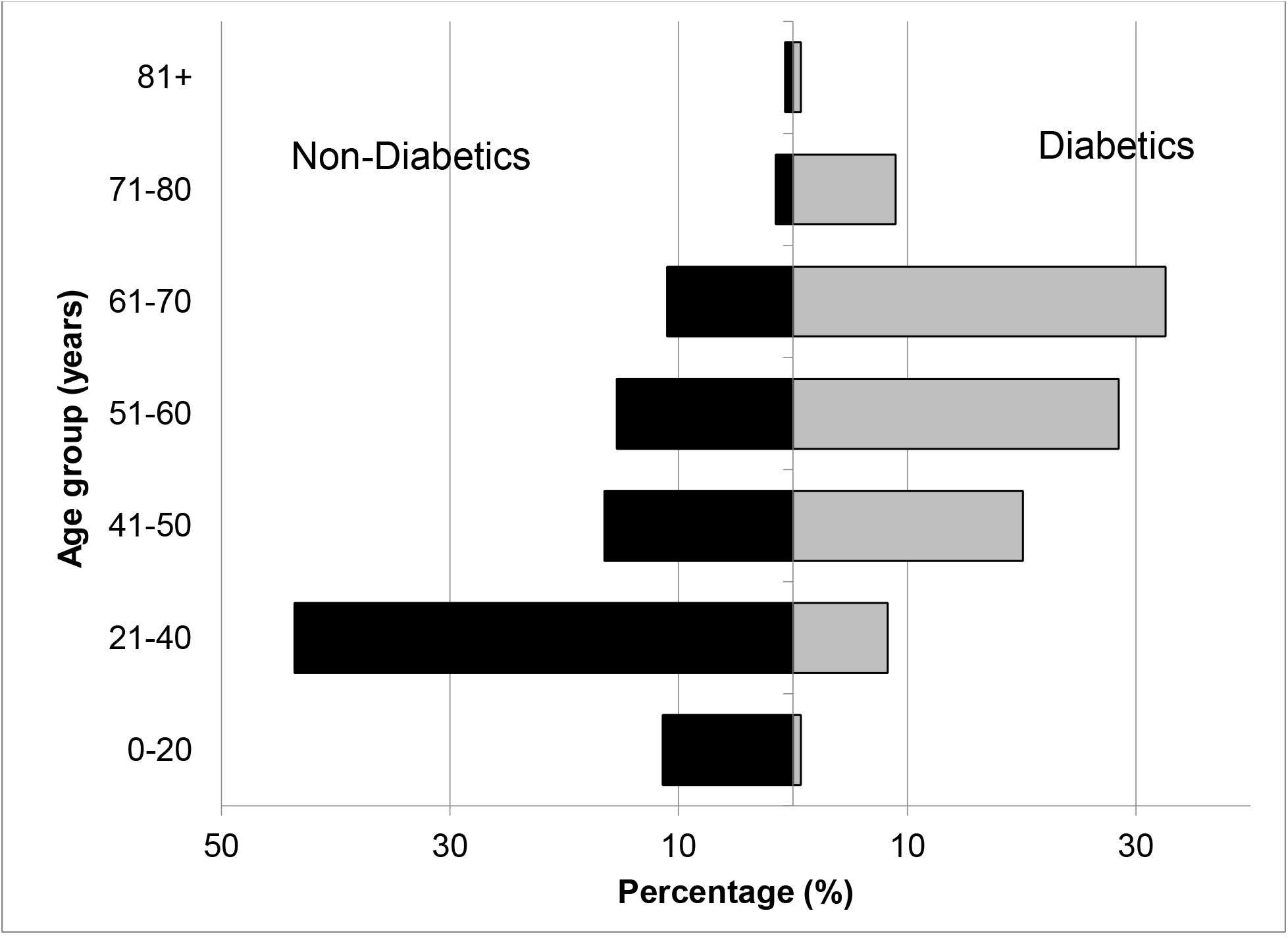
Cohort age structure stratified by diabetes status. **On average**, non-diabetic group had a mean age of 39.55 (± 16.59) years while diabetic group had a mean age of 56.44 (± 11.64) years (p-value <0.001, student’s T test).

The diabetic group had a higher prevalence of comorbidities including hypertension, asthma, cardiovascular disease, and chronic renal disease (*p* < 0.001, < 0.05, < 0.001, and < 0.005, respectively; Table 1).

### 3.2 COVID-19 severity, outcome, and associated complications

In terms of COVID-19 severity, we found no statistically significant difference in the percentages of asymptomatic cases between the diabetic and non-diabetic groups (Table 2). However, the diabetic group included a significantly higher proportion of patients requiring admission to the ICU (*p* < 0.001). Without adjustment, the diabetes group had a significantly higher percentage of death as compared with the non-diabetic group (16.7% vs. 12.1%, *p* < 0.001; Table 2). The diabetic group also had a significantly higher prevalence of complications, including sepsis, ARDS, cardiovascular disease, heart failure, and kidney injury when compared with the non-diabetic group (*p* < 0.001; Table 2).

**Table 2.** COVID-19 severity, outcome and associated complications among diabetics and non-diabetics

|  | Non-Diabetics<br>(n=273) | Diabetics<br>(n=144) | P-value |
| --- | --- | --- | --- |
| **Disease Category** |  |  |  |
| Asymptomatic | 119 (43.6) | 45 (31.3) | 0.014 |
| Mild/Moderate | 133 (48.7) | 38 (26.4) | **<0.001** |
| Severe<br>"including death<br>cases" | 21 (7.7) | 61 (42.4) | **<0.001** |
| Death cases | 10 (3.7) | 50 (34.7) | **<0.001** |
| **In-hospital complications** |  |  |  |
| Sepsis | 13/273 (4.8) | 44/144 (31) | **<0.001** |
| ARDS | 18/273 (6.6) | 61/144 (42.4) | **<0.001** |
| Heart Failure | 9/273 (3.3) | 43/144 (29.9) | **<0.001** |
| Cardiac Injury | 7/273 (2.6) | 28/144 (19.4) | **<0.001** |
| Kidney Injury | 6/273 (2.2) | 35/144 (24.3) | **<0.001** |

### 3.3 Fasting blood glucose level and odds ratio of death

We used a logistic regression model and reported the odds ratios of the outcome of dying from COVID-19 for each 1-mmol/L increase in fasting blood glucose. We found that every 1 mmol/L increase in fasting glucose is associated with 1.52 (95% CI: 1.34 – 1.72, p<0.001) times the odds of dying from COVID-19.

### 3.4 Clinical biochemistry findings

The diabetic and non-diabetic groups were subdivided into asymptomatic, mild-moderate, and severe categories. The clinical biochemistry findings were compared between the patients with and without diabetes in the subcategories of COVID-19 severity (Table 3). The cohort was categorized and divided according to the fasting blood glucose level. We noted a significant difference in fasting blood glucose between the diabetic and non-diabetic groups across all subcategories. The mean estimated glomerular filtration rate (eGFR) was significantly lower in the diabetic group than the non-diabetic group in all subcategories. The lowest value was in the severe group (57.50 ± 33.81 vs. 76.46 ± 34.14 mL/min/1.73 m^2^, *p* < 0.05). The other renal markers, creatinine and urea, were higher in general in the diabetic groups than the non-diabetic groups. However, the results were significant only in the severe subcategory (*p* < 0.05 for both markers). The C-reactive protein levels were significantly higher among diabetic patients than non-diabetic patients in all subcategories (*p* < 0.05, < 0.05, and < 0.005, in order; Table 3). Although procalcitonin levels were higher in all diabetic groups, the difference was not statistically significant. The albumin level was significantly lower in the diabetic groups across all subcategories than the non-diabetic groups (*p* < 0.001, < 0.001, and < 0.05, in order). White blood cell counts, and neutrophil counts were significantly higher in the diabetic group in the severe subcategory as compared with the non-diabetic group (*p* < 0.05 and < 0.005, respectively; Table 3).

**Table 3.**
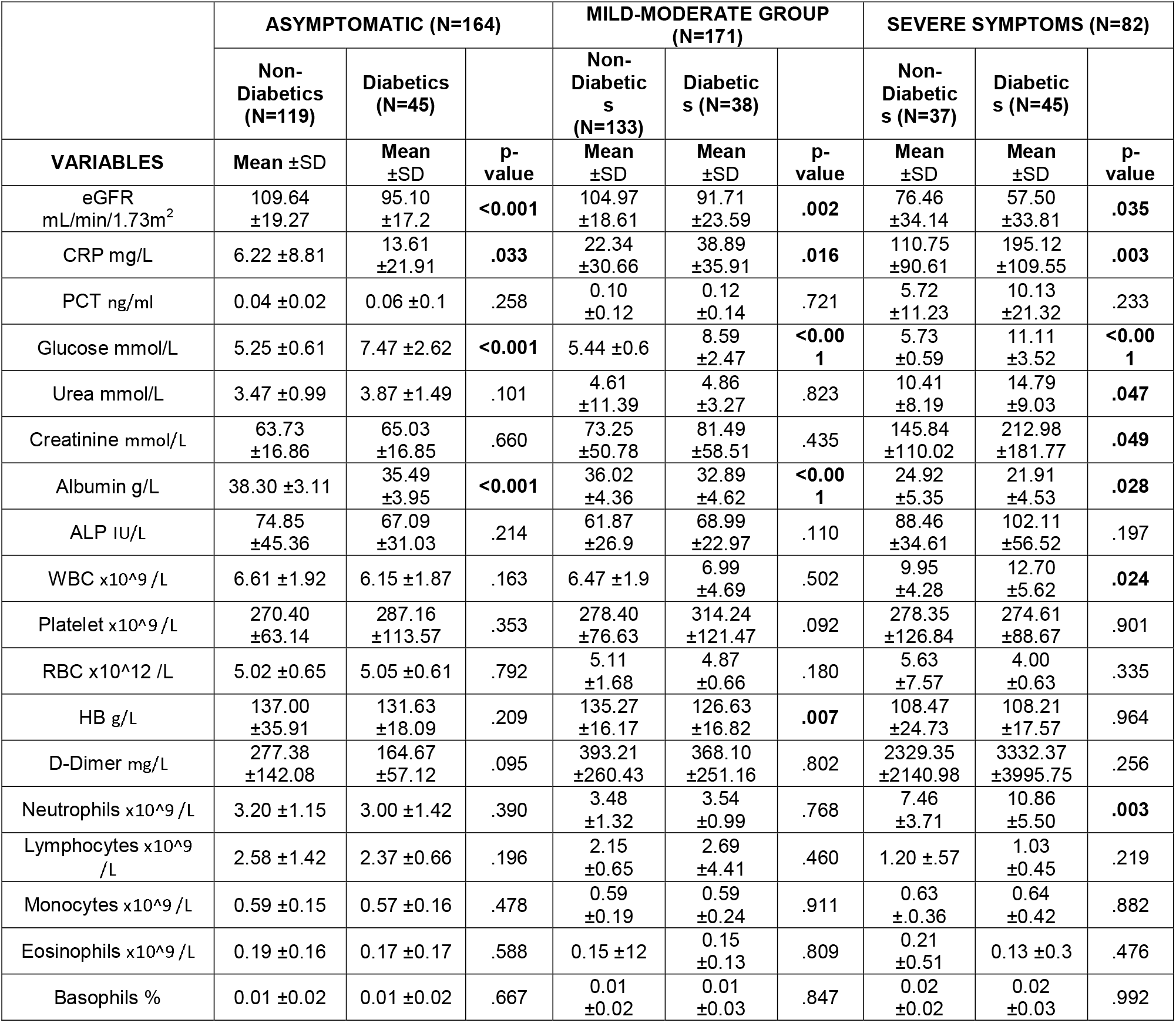
Clinical biochemistry findings per group stratified by diabetic status

## 4. Discussion

COVID-19 has displayed a broad spectrum of severity, ranging from asymptomatic to severe illness leading to death. Many factors influence disease outcome, including age and gender. Severe outcomes have been associated with preexisting chronic illnesses, such as hypertension and diabetes. In this study, we presented the clinical characteristics and outcomes of diabetic COVID-19 patients in Kuwait.

In general, we found that patients with diabetes had more severe COVID-19 outcomes than patients without diabetes, represented by the higher proportion of ICU admitted cases (severe) and deaths (Table 2). Our results are in agreement with the findings obtained from a Chinese COVID-19 cohort, in which COVID-19 diabetic patients had a 7.3% increased risk of mortality compared with 2.3% for the general population^31^. Moreover, a British cohort showed that COVID-19 patients with uncontrolled diabetes had a higher risk of death than other patients did^32^.

The impact of uncontrolled diabetes, with emphasis on hyperglycemia, continuously appears to be indicative of a poorer outcome of COVID-19 ^33^. Studies have stated that this may be linked to hyperglycemia playing a detrimental role in overproduction of interleukin-6 (IL-6), which has been associated with increased lung infiltration and severity of COVID-19 ^34^. Hence, researchers have attempted to show that the use of certain glucose lowering medications may improve outcomes of diabetic COVID-19 patients. Sardu et al, showed that in a cohort of 59 patients, 26 of whom had previously diagnosed diabetes, those given intravenous insulin had a more positive outcome than those who were not ^34,35^. However, Chen et al found that COVID-19 patients taking insulin, although they presented with significantly different inflammatory markers (higher CRP, procalcitonin and erythrocyte sedimentation rate), they did not have an overall difference in severity than to patients not taking insulin ^36^.

The reason for such differences in severity and outcomes in diabetic patients with COVID-19 is likely due to the multifactorial syndromic nature of diabetes. In our cohort, for instance, we found that diabetic patients had a higher percentage of underlying comorbidities such as hypertension and cardiovascular disease (57.6% and 14.1%, respectively), compared with the non-diabetic patients (14.7% and 4.4, % respectively; Table 1). Such results agree with the findings reported in a study conducted in Wuhan, China (56.9% and 20.9%, respectively)^37^. Hypertension has continuously been associated with a severe prognosis of COVID-19. This may be related to the role of angiotensin-converting enzyme 2 (ACE2), which is widely dispersed in the body but found in abundance in lung epithelial cells. ACE2 is also found lining the cells of the pancreatic islets, hence patients with diabetes and hypertension can be treated with ACE inhibitors (ACEi) ^38^. It has been established that SARs-CoV-2 enters the cell via the ACE2 receptor, hence the administration of ACEi to these patients can potentially exacerbate the progression of COVID-19^23^. However, studies have shown that anti-hypertensive medications appear to have no impact on the prognosis of COVID-19 ^34^.

Furthermore, we found significantly higher levels of C-reactive protein in all of our COVID-19 diabetic patients than non-diabetic COVID-19 patients (Table 3). Inflammation plays a critical role in diabetes pathogenesis, whereby diabetic patients typically develop a chronic state of inflammation^39^. Hence, this finding could be associated with an inflammatory status that makes these patients more susceptible to the damaging effects of what is known as the COVID-19 cytokine storm, which leads to multiorgan failure and death^40^.

We also noted a significantly lower eGFR upon admission for diabetic patients as compared with non-diabetic individuals (Table 3). This finding coincided with a higher incidence of acute kidney injury among diabetic patients than non-diabetic patients (24.3% and 2.2%, respectively, *p* < 0.001; Table 2). Diabetic patients have a higher risk of developing chronic kidney disease, which can eventually lead to kidney failure. The situation is worse in diabetic COVID-19 patients, because SARS-CoV-2 is likely to target the kidney through anACE2–dependent pathway, leading to renal impairment and death^41^.

Our logistic regression model showed a positive correlation between fasting blood glucose levels and increased risk of COVID-19 death, with adjustment for other risk factors including age, gender, smoking status, and other comorbidities. This finding might suggest that elevated glucose levels could be the primary molecular trigger for a cascade of pathological events that contribute to the poor outcome associated with COVID-19 in diabetic patients.

## 5. Conclusion

In conclusion, diabetes is one of the major risk factors associated with the poor outcome and mortality of COVID-19 patients. COVID-19 patients with diabetes have a higher prevalence of comorbidities such as hypertension, higher levels of inflammatory markers, lower eGFR, and a higher incidence of in-hospital complications, which illustrates the possible multifaceted pathological mechanistic pathways triggered by hyperglycemia that lead to worse outcomes and mortality.

## Data Availability

Data presented in this study was made available on springer nature data depository.

https://doi.org/10.6084/m9.figshare.12567881.v1.

## Acknowledgments

We would like to thank the administration of Jaber Al-Ahmad hospital and KFAS Emergency Resilience Program for facilitating and supporting this work. No funding was obtained for the presented work.

## Supplementary

